# Pathway-specific polygenic scores substantially increase the discovery of gene-adiposity interactions impacting liver biomarkers

**DOI:** 10.1101/2025.05.19.25327939

**Authors:** Kenneth E. Westerman, Daniel I. Chasman, W. James Gauderman, Arun Durvasula

**Author notes:** Correspondence (KEW); (AD).

## Abstract

Polygenic scores (PGS) have been leveraged to detect gene-environment interactions across many complex traits and environmental variables. While PGS×E regression is potentially more powerful than single-variant genome-wide interaction studies (GWIS) due to the aggregation of genetic effects and reduced multiple testing burden, standard PGS reflect many different biological mechanisms, limiting interpretation and potentially diluting pathway-specific interaction signals. Previous work has uncovered significant genome-wide PGS×BMI signal for liver function, but there is an opportunity for additional and more interpretable discoveries. Here, we leverage pathway-specific polygenic scores (pPGS) to discover novel mechanism-specific gene-adiposity interactions. We tested for adiposity interactions impacting three liver-related biomarkers (ALT, AST and GGT) using (1) a standard, genome-wide PGS, (2) an array of pPGS containing variant subsets derived from KEGG pathways, and (3) a GWIS. For ALT, we identified 49 significant pPGS×BMI interactions at a Bonferroni corrected *p* < 2.7×10^−4^, 80% of which were not explained by genes close to the 8 loci found in the associated GWIS. Across all biomarkers, we found interactions of BMI with 83 unique pPGS-based on KEGG pathways. We tested alternate pathway collections, including Hallmark gene sets and the KEGG Medicus database, finding that the choice of pathway collection strongly impacts discovery. Our results support the use of pPGS for well powered and interpretable discovery of pPGS×E interactions with adiposity-related exposures for liver biomarkers and motivate future studies using a broader collection of exposures and outcomes.

## Introduction

Recent work has shown that gene-environment interactions (G×E) have a substantial impact on complex disease and trait variation^1–6^. However, discovery and interpretation of single-variant GxE via genome-wide interaction studies (GWIS) is challenging due to the large number of hypothesis tests that exacerbate the intrinsically low power of interaction tests^7^. Meanwhile, polygenic approaches, such as polygenic score-by-E tests (PGS×E), may lose power by leveraging a strong and rarely satisfied assumption that main and interaction effects are proportional^6,8,9^. Recently, Durvasula et al^6^ proposed a conceptual G×E model in which environmental exposures modify genetic effects on specific pathways rather than across the entire genome, motivating G×E analyses on the pathway level as a compromise between the specificity of GWIS and the power of PGS×E tests.

Chasman and colleagues provided early proof-of-concept for this approach by conducting a data-driven clustering of disease-associated genetic loci followed by cluster-specific G×E testing, focusing on cardiovascular diseases and type 2 diabetes^10^. Gauderman et al. focused on pre-established pathway annotations, showing by simulation that the use of pathway-based PGS×E can lead to substantial increases in power over testing gwPGS×E. They also applied the approach to colorectal cancer, aggregating genome-wide significant SNPs within annotated gene sets and then testing these for interaction at the pathway level^11^. These studies underscore the potential of pathway-specific PGS (pPGS) approaches for increasing power to detect G×E interactions, but it remains unclear how these pPGS interactions compare to the equivalent genome-wide, variant-specific interaction study and whether the results differ substantially by choice of pathway database.

Leveraging the annotation-based pathway approach, we sought to explore genetic modification of the relationship between adiposity (as measured by body mass index [BMI]) and liver stress biomarkers. Our prior work demonstrated the presence of a polygenic signature modifying the relationship between BMI and cardiometabolic risk factors^12^. There was a particularly strong signature for liver-related biomarkers, and we showed preliminary evidence that interactions with BMI reflected a mechanism-specific subset of the overall genetic architecture of these biomarkers. Here, we explore this biological question further as an applied setting to test the incorporation of existing biological annotations and pPGS into an interaction testing framework.

## Results

Our analysis pipeline is summarized in **Fig. 1**. We analyzed individual-level data for 344,000 unrelated participants of European ancestry from the UK Biobank. We performed a standard GWAS for each of three log-transformed liver-related biomarkers (alanine aminotransferase [ALT], aspartate aminotransferase [AST], and gamma-glutamyl transferase [GGT]) in the entire UKB dataset. We used these summary statistics to create a genome-wide PGS (gwPGS) as well as a series of pPGS based on KEGG pathway annotations. We then tested each PGS for interaction with BMI. For comparison to the PGS-based interaction tests, we performed a genome-wide interaction study (GWIS) for each biomarker with BMI as the exposure.

**Figure 1.**
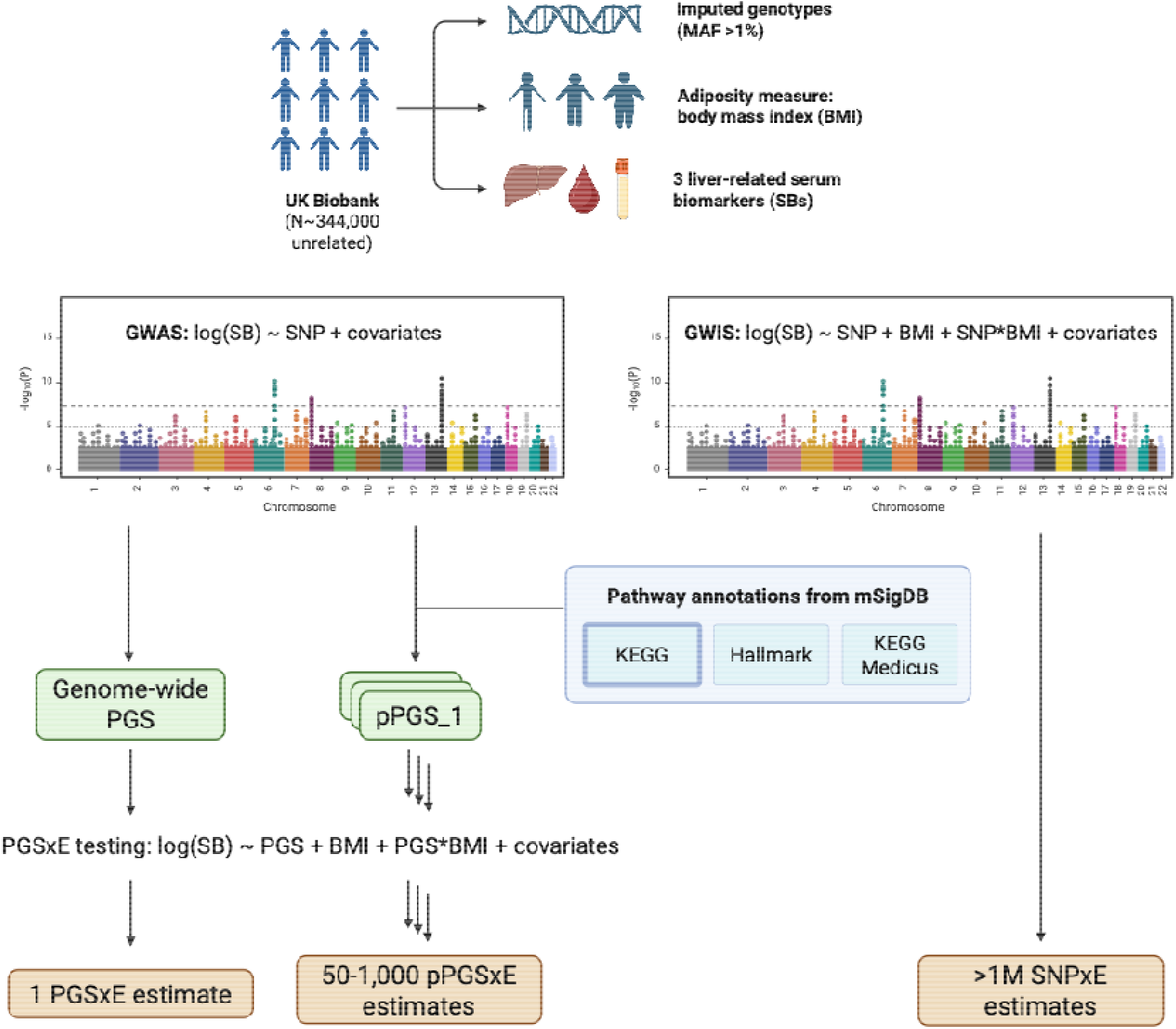
Pipeline for multi-scale gene-by-body mass index interaction (G×BMI) testing carried out in this study.

We first tested for the interaction between genome-wide PGS and BMI in a regression including a main effect for the gwPGS and adjusting for age, age^2^, sex, age×sex, 10 genomic principal components (gPCs), and 10 BMI×gPC product terms. We used a pruning and thresholding (P&T) approach to train the gwPGS, retaining between 4612 and 6164 SNPs at a *p*-value threshold of 0.001. We identified strong gwPGS×E interactions for all three liver biomarkers. For ALT, AST, and GGT, these interaction effect sizes were 0.035 SD_log(ALT)_ SD_PGS_ SD_BMI_ (*p* = 1.7×10^−133^), 0.028 (*p* = 7.8×10^−74^), and 0.022 (*p* = 3.7×10^−58^), respectively. These results are concordant with our recent analyses of PGS×BMI for liver biomarkers^12^, and all represent patterns in which higher PGS values magnify the positive BMI-biomarker relationship. We note that there is no expected bias due to training a standard PGS and testing it for interaction in the same dataset, as reported by Gauderman and colleagues^11^.

Next, we investigated the discovery enabled by pathway-level testing. We assigned SNPs to genes based on physical distance to the closest annotated gene (−2kb to +1kb from the gene transcription start site and transcription end site, respectively) and assigned genes to pathways using gene sets from the mSigDB database, focusing on the KEGG gene-to-pathway mappings as the primary pathway group of interest (**Fig. S1**). We calculated pPGS for each pathway using a *p*-value-based pruning and thresholding (P&T) approach as implemented in the PRSet program^13^, using a *p*-value threshold of 0.001. We tested for interactions between pPGS and BMI using regression models analogous to those used for the standard gwPGS above, assigning significance based on a Bonferroni correction for the 186 total pathways tested. We identified a substantial number of significant pPGSxE for all three biomarkers (**Fig. 2b** for ALT; full set of results in **Table S1**). First, we found 49 significant pPGSxE interactions for ALT. Though no single pPGS reached the same degree of significance as the gwPGS, interactions were highly significant for multiple pathways, including glycerolipid metabolism (*p* = 2.0×10^−122^) and focal adhesion (*p* = 2.9×10^−60^) (**Fig. 2a**). The glycerolipid metabolism association is concordant with previous work showing a relationship between glycerolipid metabolism and obesity^14^ as well as obesity and liver biomarkers^15^. Results for AST were similar, with the same top two pathways and 37 significant pathways in total (**Fig. S2**). Notably, for AST, the same glycerolipid metabolism pathway reached a greater degree of significance than the gwPGS (*p* = 2.1×10^−133^), highlighting the value of the pPGS in improving power by prioritizing relevant genomic regions. GGT analyses resulted in 31 significant pathways, with a notably different pattern of significance across pathways compared to ALT and AST, suggesting a different architecture of GxEs (**Fig. S3**). Here, the top pathways included regulation of actin cytoskeleton, pathways in cancer, peroxisome, and glutathione metabolism. We found models that included all pPGS×BMI interactions effects fit the data significantly better than models with only gwPGS×BMI interaction effects (ALT *p*=5×10^−66^, AST *p*=4×10^−99^, GGT *p*=1×10^−7^; see Methods for description of likelihood ratio tests). Taken together, these results show that pPGS can break down genome-wide signals of PGSxE into biologically interpretable results.

**Figure 2.**
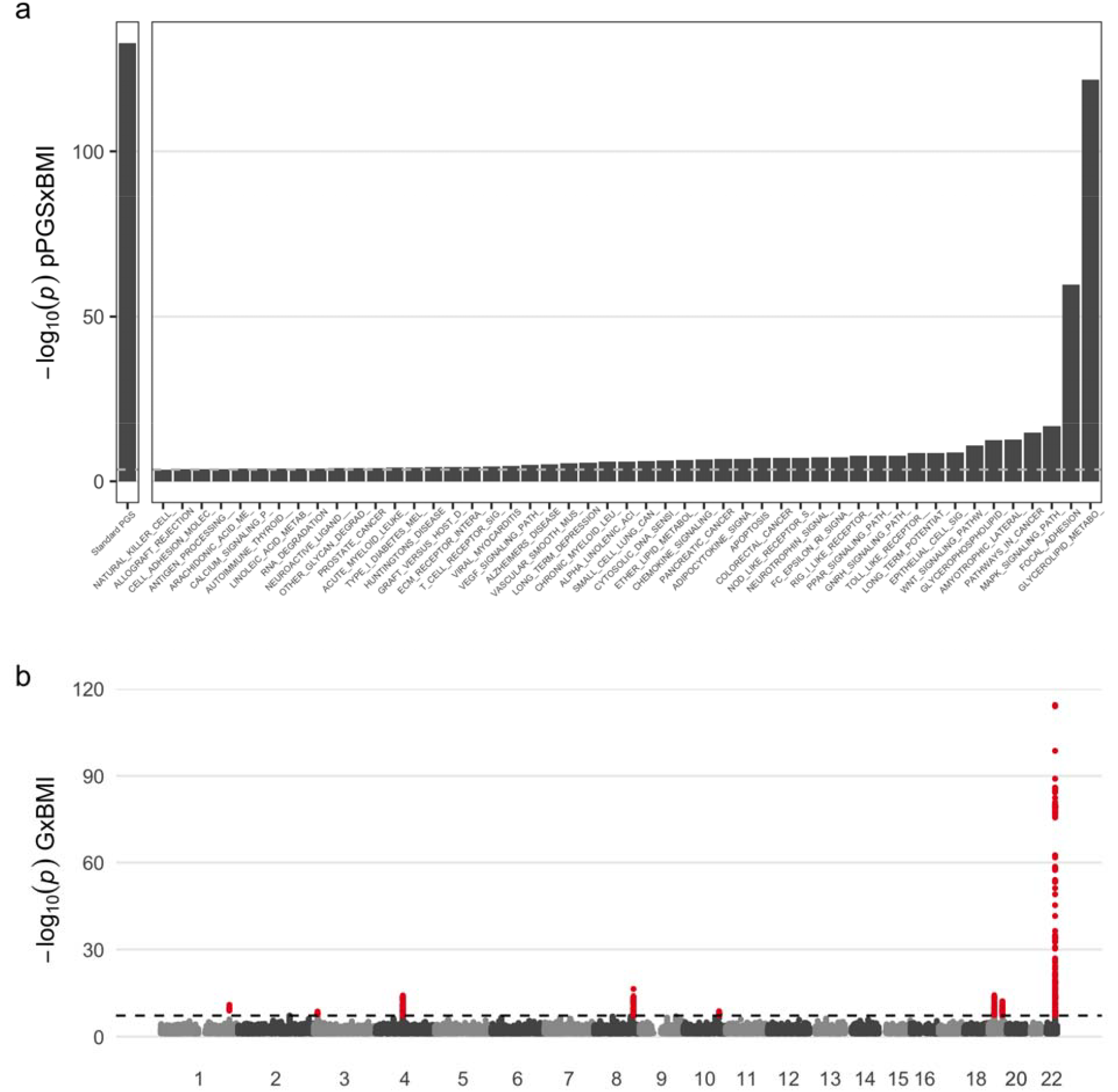
GxEs shape the relationship between adiposity and alanine aminotransferase (ALT). (a) PGSxE regression *p*-values for the gwPGS (left panel) and each significant pPGS (right panel). (b) Manhattan plot shows SNP-specific *p*-values as a function of genomic position. Results for AST and GGT are provided in **Figures S2-3**. Numeric results for all biomarkers are provided in **Tables S1-2**.

Next, we performed single-variant GWIS for each biomarker using analogous regression models to that for the gwPGS and pPGS, additionally using robust standard errors and applying a genome-wide significance threshold of 5×10^−8^. We found 8 genome-wide significant loci for each of ALT and AST (7 of these were overlapping) and 2 for GGT (**Fig. 1b, S2, S3, Table S2**) after a simple distance-based pruning (+ - 500kb from lead variants). The strongest locus for both ALT and AST mapped to the *PNPLA3* gene on chromosome 22, a well-known locus affecting liver disease and metabolic traits^16^, whose strong main effect on ALT and AST has previously been shown to be modified by BMI in UKB^17^. All 8 genome-wide significant GWIS loci also achieved significance in the associated GWAS for liver biomarkers, in analysis examining solely main effects.

Next, we asked how many of the pPGSxE results overlapped with biological pathways that could have been discovered via the GWIS approach. We annotated pPGSxE tests as “explained” if there was at least one GWIS lead variant near any gene in that pathway set (within 100kb of the TSS based on Ensembl GRCh37 v75). For ALT, AST, and GGT, we found that 39 (80%), 34 (92%), and 22 (71%) pathway interactions were unexplained by significant GWIS loci (**Table 1**), respectively. These findings indicate that the majority of pPGS×E interactions point to pathways that would not have been found using a single-variant approach. When testing for enrichment of GWIS signal using the MAGMA tool and the same set of variant-gene-pathway annotations, we found that results were significantly correlated with those from the pPGS×E approach (*p*_*Spearman*_ = 0.25 for -log_10_(*p*) values, *p* = 2.8×10^−9^), but systematically less significant (**Fig. S4**). In addition, all pathways highlighted by this GWIS enrichment approach were discovered by the pPGS×E approach. The two strongest pathways from the pPGS×E approach for ALT and AST (glycerolipid metabolism and focal adhesion) showed minimal signal in the enrichment-based approach (p > 0.05 for both). Taken together, these results highlight the increased power of pPGS×E to discover G×E interactions over GWIS.

**Table 1.** Overlap between significant findings from the single-variant GWIS and pPGS approaches.

| Biomarker | # GWIS loci | # pPGS×BMI | # "unexplained" pPGS×BMI pathways | Fraction "unexplained" pPGS×BMI pathways |
| --- | --- | --- | --- | --- |
| ALT | 8 | 49 | 39 | 80% |
| AST | 8 | 37 | 34 | 92% |
| GGT | 2 | 31 | 22 | 71% |

To understand the impact of the choice of pathway database (i.e., gene-to-pathway mapping) on our results, we reran the same analysis pipeline using two additional pathway collections from mSigDB: hallmark pathways and KEGG Medicus (see **Fig. S1** for metadata describing these collections). Over all biomarkers, these collections resulted in 83 (KEGG), 36 (hallmark), and 67 (KEGG Medicus) significant PGSxBMI interactions. The comparison of KEGG Medicus, an expansion of KEGG to include disease- and drug-related annotations, to the primary KEGG “legacy” collection indicates the importance of pathway collection choice: fewer significant pathways were uncovered despite a much larger number of available pathways (658 for Medicus versus 186 for legacy) due to the specific biological pathways represented and the differential multiple testing burden.

We used a series of secondary analyses to address potential concerns. First, we chose a single P&T *p*-value threshold of 0.001 based on optimized main effect PGS thresholds from a prior UKB analysis^12^. We confirmed that this choice did not substantially impact the pPGS×E results: interaction effects based on pPGS using a P&T threshold of 5×10^−8^ showed minimal difference in significance (**Fig. S5**). Second, we calculated an adjusted (or “effective”) number of pathways discovered to account for the fact that some pathways are correlated (see Methods for details on the PCA-based method). This resulted in a reduction of significant pathways from 49 to 9.6 for ALT, 21 to 11.1 for AST, and 10 to 2.8 for GGT when using the KEGG legacy pathways.

## Discussion

We leveraged pathway annotations and the UK Biobank with complex trait, exposure, and genetic data to identify gene-environment interactions acting on specific pathways. We found 83 significant KEGG pathway pPGS×BMI interactions across three liver biomarkers, pointing to specific biological processes modifying the relationship between adiposity and liver health.

The highly significant glycerolipid pathway finding for ALT and AST captures an interaction that we and others have described: genetic effects on this pathway’s function alter the liver’s ability to relieve adiposity-associated lipid buildup^12^. The top hit for GGT was regulation of actin cytoskeleton; this may point to the importance of cytoskeletal and mechanical integrity for maintaining proper bile flow, whose deterioration impacts GGT more than ALT or AST^18^.

Obesity-associated lipid buildup can increase mechanical stress in hepatocytes^19^ and disrupt the bile canaliculus network^20^, with potential modification of this effect by genetic effects on cytoskeletal integrity. While the preceding pathways have been studied in the context of liver health, additional highly significant pathways from the pPGS interaction approach may point to novel mechanisms of obesity-related liver pathogenesis. For example, the disease-associated amyotrophic lateral sclerosis (ALS) pathway was one of the strongest for ALT. While ALS is a motor neuron disease not classically associated with liver dysfunction, patients are highly enriched for hepatic steatosis, possibly via mitochondrial and endoplasmic reticulum stress pathways^21^, suggesting directions for further study.

Our study represents an advance over previous studies investigating pathway-specific polygenic scores for interaction. Our approach based on *a priori* pathway annotations increases the interpretability of our pPGS×E results compared to data-driven approaches. Leveraging these strengths, we show that pPGS×E testing enables increased discovery of gene-environment interactions and produces interactions that (1) are mostly unexplained by the associated variant-specific GWIS and (2) can be stronger than the associated gwPGS×E test. Furthermore, we show that the choice of pathway annotation (variant to gene to pathway) has a substantial impact on pPGS×E results and more comprehensive exploration of additional pathway collections is likely to further increase the number of interactions uncovered.

Our study has several limitations. First, we used a simple distance-based method for variant-to-gene mapping and a limited set of pathways annotations. Future work could apply more sophisticated and wide-ranging methods making use of functional genomic data to assign SNPs to genes and ultimately pathways. Second, we study a limited subset of biomarkers and only one exposure. Our methodological conclusions may not hold universally across all biomarkers and exposures. Third, we did not account for correlated pathway annotations, which makes our stringent Bonferroni correction for the total number of pathways conservative. Future work could model the correlated tests to increase statistical power. Despite these limitations, our work highlights pathway-specific PGS×E testing as a powerful way to discover G×Es.

## Methods

### UK Biobank data

We used data from the large, prospective UK Biobank cohort in all analyses^22^. This research was conducted using the UK Biobank resource under application no. 277892 and Not Human Subjects Research determination NHSR-4298 at the Broad Institute of MIT and Harvard. Genotyping, imputation, and initial quality control on the genetic dataset have been described previously^23^. Work was conducted on genetic data release version 3, with imputation to both Haplotype Reference Consortium^24^ and 1000 Genomes Project (1KGP)^25^. Ultimately, analysis was performed on a set of unrelated individuals, defined as the set of individuals whose genomes were included in centrally performed genetic principal components analysis. As described in prior related work^12^, we excluded individuals that had withdrawn consent by the time of analysis excluded as well as those with diabetes, coronary heart disease, cirrhosis, end-stage renal disease, cancer diagnosis within one year prior to their assessment center visit, or who were pregnant within one year of the assessment center visit.

Body mass index (BMI; kg m^2^) was collected from assessment center anthropometric measurements. Serum ALT, AST, and GGT values were measured in blood samples collected at the baseline visit (details available at: https:biobank.ctsu.ox.ac.ukcrystalcrystaldocsserum_biochemistry.pdf), and each were log-transformed prior to analysis.

### Genome-wide studies

A genome-wide association study (GWAS) and genome-wide interaction study (GWIS) was performed for each log-transformed biomarker. The GWAS used a basic linear model corrected for covariates:

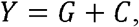

We adjusted for the following covariates: age, age^2^, sex, and ten genetic PCs. We used model-based standard errors and included only variants with minor allele frequency >1% and imputation quality INFO score >0.5 (approximately 9,891,000 variants in total). The GWIS model added an environmental exposure and its product term with G:

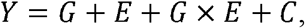

In the GWIS, we additionally adjusted for an E×gPC product term for each gPC^26^ and used robust standard errors. All genome-wide studies were conducted using GEM v1.5.2^27^.

### Pathway annotations and polygenic score generation

We generated PGS from GWAS summary statistics using the PRSet program^13^, which builds on PRSice-2^28^. PRSet both computes pathway-specific PGS weights (using the P&T approach along with an LD reference panel) and calculates scores for the input UKB dataset as a linear combination of genotypes based on those weights. As input parameters governing the behavior of the P&T algorithm, we used *p*-value thresholds of both 0.001 and 5×10^−8^, a clumping radius of 1MB, an r^2^ threshold of 0.1, and an LD reference panel consisting of a random 20,000 individuals from the UKB. In all analyses, we considered only autosomal variants and removed ambiguous variants (A T or C G) during PGS development.

To generate pPGS, we additionally obtained pathway annotations from mSigDB^29^. We used three sets of pathway collections: 1) KEGG^30^, 2) Hallmark^29^, and 3) KEGG Medicus^30^. PRSet assigns variants to pathways via physical proximity to constituent genes and subsequently conducts a separate P&T procedure for each pathway. For this variant to gene mapping, we included variants within a boundary of 2kb upstream (5’) and 1kb downstream (3’) of the gene transcription start site and end site, respectively. By default, PRSet also generates a gwPGS using the same P&T procedure but including all available variants, resulting in gwPGS containing 4,612, 4,894, and 6,164 variants for ALT, AST, and GGT, respectively.

### Polygenic score by environment testing

We tested for interactions using linear regression of the form:

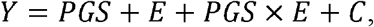

where Y is the trait value, PGS is the polygenic score for the trait, E is the environment variable, and C is a set of covariates. For pathway PGS, we used the following linear regression for a single pathway:

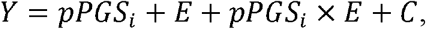

Where *i* ∈ *P* indexes the pathway in the set of pathways P (see Pathway annotations and polygenic score generation). For all analyses, we correct for the same covariates as in the GWIS: age, age^2^, sex, ten genetic PCs, and an E×gPC product term for each gPC^26^. We used the p-value associated with the interaction term to assess significance. To account for multiple testing in the pathway specific PGS, we used a Bonferroni corrected threshold of 0.05 divided by the number of pathways tested. All individual-level data preprocessing and regression analysis after PGS generation was performed using R v4.1 and 4.2 ^31^ except where otherwise noted.

We used likelihood ratio tests to understand whether the cumulative contribution of many pPGS×BMI interactions explained significantly more variance than the gwPGS×BMI interaction alone. The restricted model included all covariates from the basic pPGS×BMI interaction tests (see above) as well as a main effect for the gwPGS, main effects for each pPGS, and an interaction term for the gwPGS. The full model added interaction terms for each of the pPGS. We tested for significance of the additional variance explained by the full model using the *lmtest::lrtest()* function.

To enable us to a report a number of discoveries that accounted for the substantial correlation between pPGS, we calculated an “effective” number of pathways discovered for each biomarker using a previously described method^1^. Briefly, we subsetted a rectangular matrix of individual-level pPGS values to include only those that were significant for the biomarker of interest, then performed principal components analysis (*prcomp* function with standardized variables). The number of effective biomarkers was then calculated from the principal component variances, *λ* (equal to the eigenvalues of the biomarker covariance matrix) as 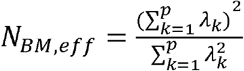.

## Data availability

No new genetic or phenotypic data have been generated for this study. The UK Biobank data, including genetic and phenotypic data, are under controlled access but can be obtained through application at https:ww.ukbiobank.ac.uk. UK Biobank will consider data applications from bona fide researchers for health-related research that is in the public interest.

