## Supplementary Figures for "Pathway-specific polygenic scores substantially increase the discovery of gene-adiposity interactions impacting liver biomarkers"

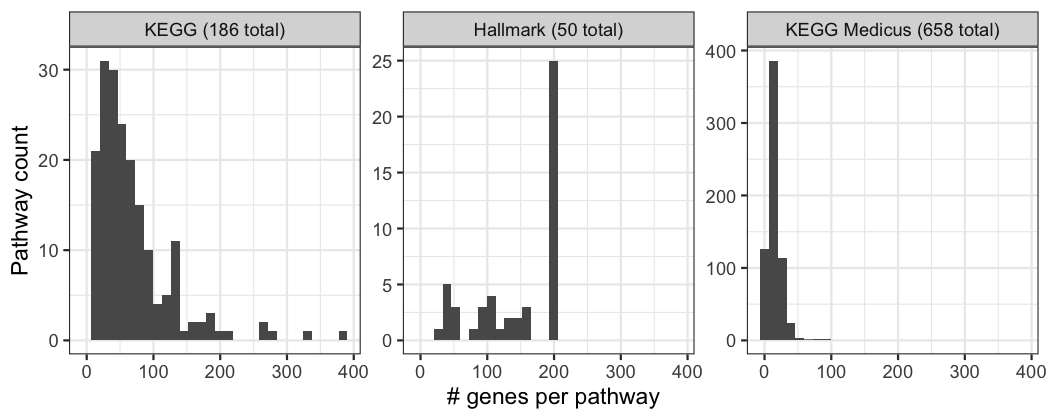
**Supplementary Figure S1.** Summary of selected pathway annotations from mSigDB. For each panel (corresponding to pathway groups, along with their total number of pathways), the histogram displays counts of pathways (*y*-axis) having a given number of constituent genes (*x*-axis).

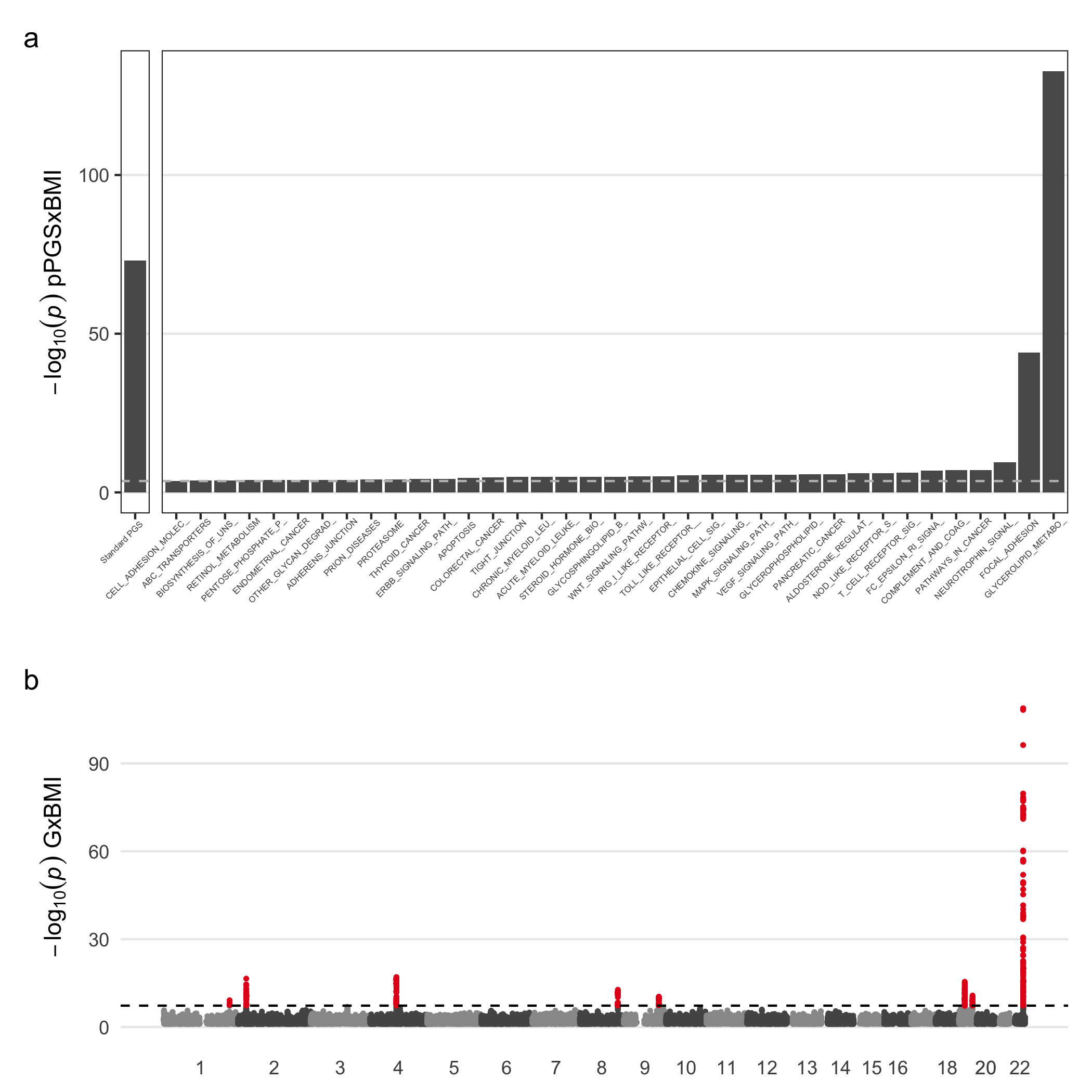
**Supplementary Figure S2.** GxEs shape the relationship between aspartate aminotransferase (AST) and adiposity. (a) PGSxE regression *p*-values for the gwPGS (left panel) and each significant pPGS (right panel). (b) Manhattan plot shows variant-specific *p*-values as a function of genomic position.

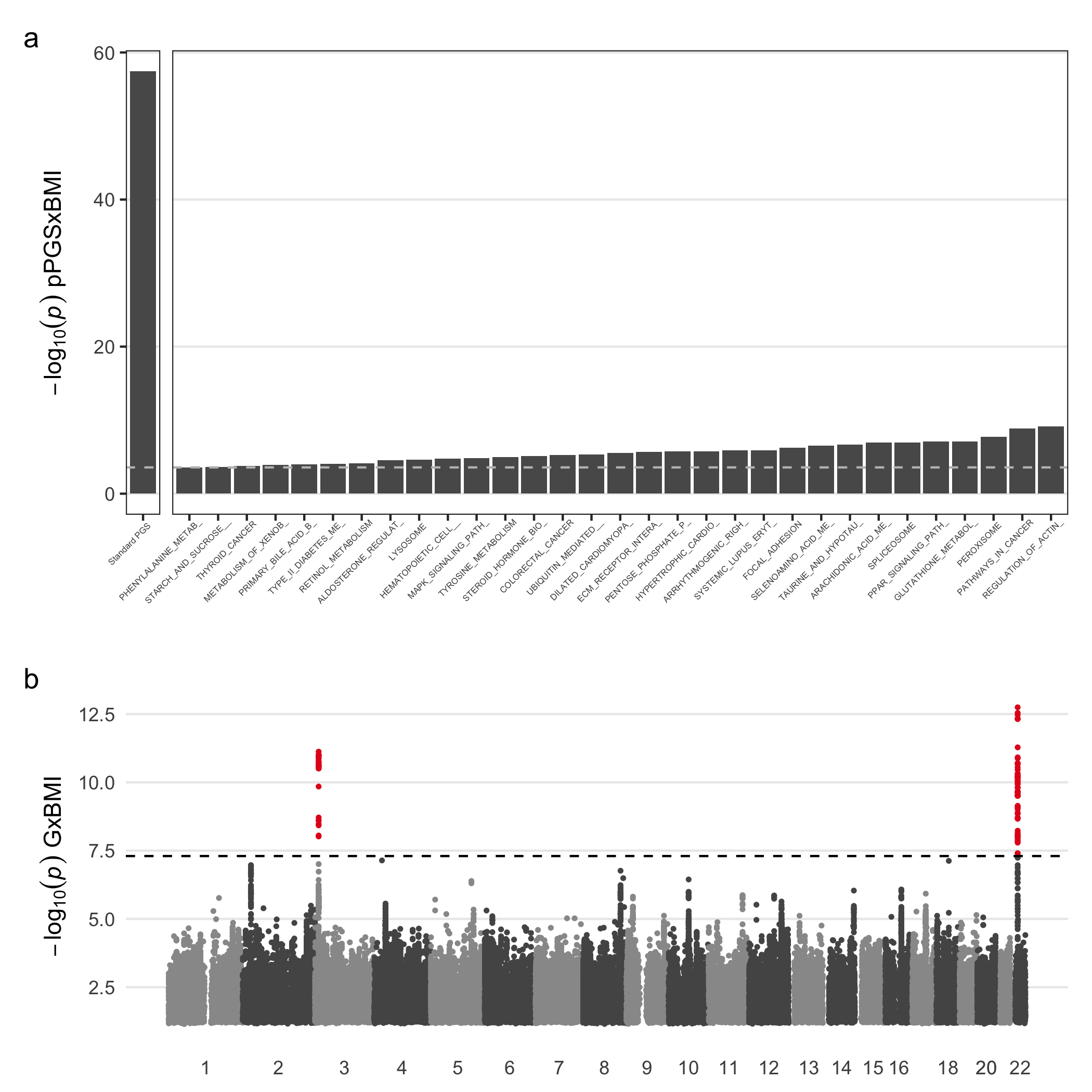
**Supplementary Figure S3.** GxEs shape the relationship between gamma-glutamyl transferase (GGT) and adiposity. (a) PGSxE regression *p*-values for the gwPGS (left panel) and each significant pPGS (right panel). (b) Manhattan plot shows variant-specific *p*-values as a function of genomic position.

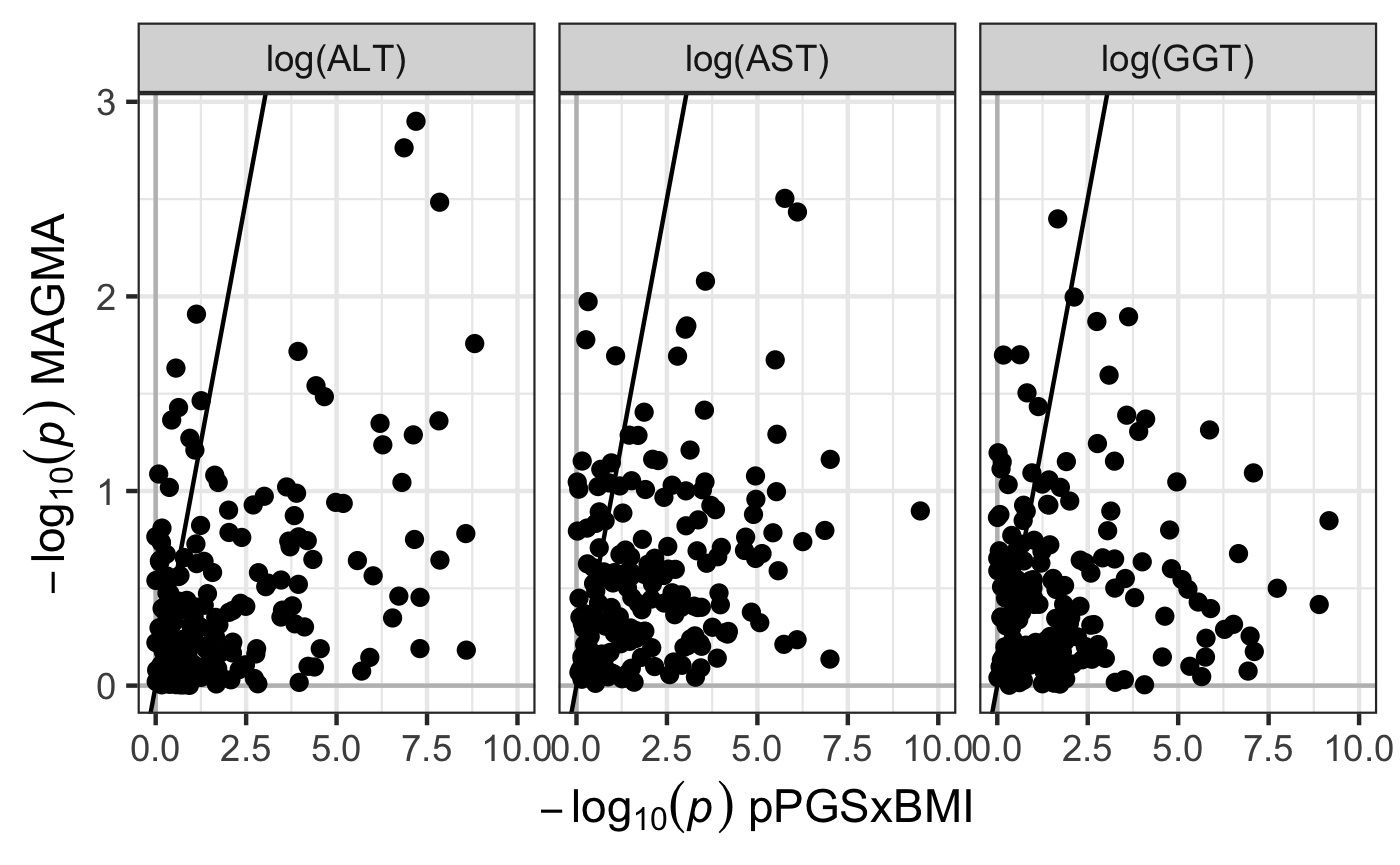
 **Supplementary Figure S4.** Comparison of the statistical significance of pathway-level interactions tests using either pPGSxE (*x*-axis) or GWIS enrichment (based on the MAGMA tool; *y*-axis). Each point corresponds to a single KEGG legacy pathway. *x*-axis has been constrained for ALT and AST to aid visualization, which excludes results for glycerolipid metabolism and focal adhesion pathways (*p*-values reported in main text). Two points (corresponding to the “Glycerolipid Metabolism” and “Focal Adhesion” pathways) have been removed from the ALT and AST plots due to their extreme values (see Results text for their *p*-values).

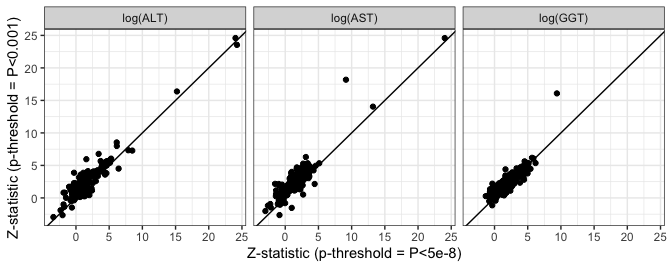
**Supplementary Figure S5.** Comparison of the statistical significance of pPGSxBMI tests using pPGS derived using a P&T *p*-value threshold of 0.001 (*y*-axis) versus 5x10^-8^ (*x*-axis). Each point corresponds to a single KEGG legacy pathway. We note that points could not be plotted for pathways having no annotated variants that pass *p*<5x10^-8^.
